# Profiling ECD and related data systems and key lessons in Kenya and across Sub-Saharan Africa: a protocol of a scoping review

**DOI:** 10.1101/2025.09.11.25335595

**Authors:** Keneth Okello, Evitar Aluoch Ochieng’, Daniel Mwanga, Eubert Munywere, Nelson Langat, Patricia Kitsao-Wekulo, Linda Oloo, Paul Otwate, Calistus Wilunda, Amanuel Abajobir, Silas Onyango, Charity Waweru-Mwangi, Margaret Nampijja

## Abstract

**Introduction:** Holistic child growth and development require a nurturing environment that enhances the well-being of the child in all aspects of their life (health, growth, development). However, there is currently a lack of aggregated information and data on how children thrive across the five domains of the nurturing care framework (NCF), especially in communities facing multiple vulnerabilities or adversities. This scoping review aims to profile existing ECD and related data systems in sub-Saharan Africa (SSA), with a particular focus on Kenya’s ECD data systems and an even deeper profiling in three counties (Nairobi, Isiolo and Homa Bay). This paper describes the protocol of the scoping review.

**Methods:** This scoping review protocol follows guidelines by Levac et al. (Levac et al., 2010) and builds on the scoping review framework proposed by Arksey and O’Malley (Arksey & and O’Malley, 2005). This review is expected to begin in October 2025. The study team will search peer-reviewed literature from databases including PubMed, ScienceDirect, African Journals Online (AJOL), Scopus, APA PsycINFO, and PsycNet, covering the period from October 2015 to September 2025. A grey literature search will also be conducted using platforms such as Google Scholar, OpenGrey, and national and sub-national websites and reports. The search will be performed using a list of search terms that are expected to generate relevant literature. Relevant literature will be reviewed, and data will be extracted. Relevant information will be organised into study characteristics and key details, including the types of ECD and related data systems and platforms, which ECD indicators they used and their sources, which other systems they were linked to, which stakeholders were involved, and the lessons and experiences, including what worked and the challenges, will be highlighted.

**Ethics and dissemination:** No ethical approvals are required for this scoping review because the data will be gathered from existing published literature, and no new data will be collected. Nevertheless, the project within which the scoping review is part received ethics approval from AMREF and NACOSTI in Kenya. The findings of the review will be used to plan the subsequent phases of our project and shared with stakeholders through workshops, publications in peer-reviewed journals, and conferences.

**Protocol Registration:** This protocol has been registered at OSF on 4^th^ April 2025. https://doi.org/10.17605/OSF.IO/HT24X

## INTRODUCTION

About 250 million children worldwide are at risk of not achieving their developmental potential due to exposure to poverty, diseases, malnutrition, and lack of stimulating environments (Grantham-McGregor et al., 2007). The risk of failure to thrive is disproportionately higher in low- and middle-income countries (LMICs), particularly in sub-Saharan African countries where these adversities are concentrated (Gil et al., 2020). For instance, according to the 2020 World Bank data, 56% of Kenya’s urban population lives in informal settlements, which are characterized by a lack of land/tenancy rights, poor amenities, health and education services, and a high rate of unemployment and crime (APHRC, 2018). Together these circumstances result in adverse effects on the development of the world’s poorest children. Similarly, remote, hard-to-reach rural communities often grapple with unique challenges, including poverty, poor health, and malnutrition, which are consequent to prolonged droughts and their effects on child health, nutrition, and development. These communities continue to suffer from inequitable access to services, including interventions specific to early childhood development (ECD). According to the 2016 estimates, 38.3% of Kenyan children aged 3 and 4 are not reaching their cognitive and socioemotional milestones and potential (Harvard School of Public Health, 2016). A recent national ECD survey in Kenya revealed that about 25% of children were developmentally delayed (UNICEF, 2014). In Isiolo and other remote, climatically challenged frontier counties, despite 85 agricultural projects, food insecurity persists, with 28% of children malnourished (unpublished Nawiri project data). The COVID-19 pandemic has caused a rise in teenage pregnancies, especially in underserved areas like Homa Bay, which has a 24% prevalence rate (UNICEF, 2020), making it a high-risk zone for children’s development and substandard care. Early childhood adverse exposures are likely to have long-term effects on children’s later development and success if not addressed early. This poses significant negative consequences for community growth and prospects. While child survival is essential, it’s equally important that children thrive and reach their full developmental potential, as this ultimately determines their later success and well-being. Creating a nurturing environment that promotes health, growth, development, and holistic well-being is therefore critical.

However, there is currently a critical lack of aggregated information or data on how children thrive in the nurturing care framework (NCF) domains, especially in communities facing multiple vulnerabilities or adversities. Children living in areas affected by socio-economic, climatic adversities, and systemic barriers are at high risk of poor health, growth and development outcomes. However, they remain largely invisible in existing data systems. Across sub-Saharan Africa, the absence of comprehensive, real-time, and actionable data limits the ability of decision-makers to implement timely, evidence-based interventions that could prevent developmental setbacks in the short-term and long-term.

Common system barriers to data on ECD services and care include inadequate financial resources, poor allocation of limited funds, and fragmented reporting mechanisms resulting from a lack of evidence on ECD indicators across the NCF domains (Health, Nutrition, Early Learning, Responsive Caregiving, Safety, and Security). This is complicated by the ambiguity surrounding the organizational structure responsible for overseeing ECD, leading to a lack of coordination and collaboration between the different sectors. Furthermore, there is a lack of a dedicated budget for ECD research and care, with funding being indicator- or sector-specific and not considering ECD needs in its entirety or relative needs between two or more aspects of ECD. The primary barrier underlying system issues is the absence of a coordinated ECD data visualization system or platform to provide aggregated data for monitoring performance and decision-making, including policies, budget allocation, and service delivery, especially for planning care for the most vulnerable children. It is on this background that we aim to conduct a scoping review, to profile the existing ECD data systems in sub-Saharan Africa (SSA) with a deeper dive in Kenya and special interest in three counties (Nairobi, Isiolo and Homa Bay) where an initiative aimed at developing an ECD data visualization system is about to be conducted. This scoping review also aims to identify key stakeholders and personnel involved in the different data systems, including their use, management, updating and maintenance. Further, we identify the specific child indicators that are captured, the tools used, infrastructure and capacity, how the data are handled, used, and maintained, which programs/software are used, and the data quality, as well as key lessons, challenges experienced and recommendations that can be used in the development of the ECD data visualization platforms for Kenya. The current paper describes the protocol of the scoping review.

## METHODOLOGY

The scoping review protocol follows the guideline developed by Levac et al. (Levac et al., 2010) by advancing the scoping review framework proposed by Arksey and O’Malley (Arksey & and O’Malley, 2005). We will follow the following stages: identifying the research question, identifying relevant studies, selecting studies, charting the data, collating, summarizing, reporting the results, and consulting.

We will conduct a rapid scoping review to map out ECD data systems in the respective counties. This will be combined with the mapping of stakeholders such as ECD technical staff, statisticians, planning officers, research and development officers, experts and consultants, and partners involved in managing and using data systems. We will assess several key areas, including the capture of specific indicators, tools, infrastructure, and capacity; the management, use, and maintenance of data; the programs and software utilised; and the overall data quality. The African Population and Health Research Center (APHRC) has previously supported countries to profile data systems for data on youth and children at national and subnational levels (APHRC, 2018). We will also conduct a scoping review of ECD data visualization platforms used in Kenya and other LMICs (e.g. Tanzania, which has made some steps towards developing an ECD dashboard and scorecard), how they work, and how they can be adapted to our context. The findings from this phase will inform the next phase, in which we will engage stakeholders in health and other sectors involved in ECD in interviews to understand the existing ECD monitoring systems within health and other sectors and any gaps (including capacity).

### Identification of the research question

The scoping reviews aim to address the following main research question: What are the existing ECD data systems, both local, national, and regional, and which stakeholders are involved in ECD data management at each level of government? Given the specificity of the study setting, we will use the 3WH framework by Barroso (9), which is what (topic), who (population), when (temporal), and how (methodology) to refine our research questions, as shown in Table 1.

**Table 1:** Research questions developed based on the 3WH (What, Who, When, How) framework.

| **What (topic)** | **Who (population)** | **When (temporal)** | **How (methodological)** | **Location/County** | **Research question** |
| --- | --- | --- | --- | --- | --- |
| Sources of ECD-related data | Under five | In the past 10 years<br>(01/10/2015 to 30/09/2025) | Academic and grey literature | Regionally, Kenya and county-specific | 1. What are the key sources of ECD-related data in the region, specifically in Kenya and Nairobi, Isiolo, and Homa Bay counties? |
| Types of data systems that currently exist and are being used | Under five | In the past 10 years<br>(01/10/2015 to 30/09/2025) | Academic and grey literature | Regionally, Kenya and county-specific | 2. What data systems exist and are used in the three target counties (Nairobi, Isiolo, and Homa Bay) for capturing/recording and reporting ECD and related data?<br>3. What electronic data systems/ platforms currently capture and report ECD indicator data in your sector (health and nutrition, education, child protection, etc)?<br>4. What level of accessibility does the ECD data system provide?<br>5. Which platforms are used to manage ECD data?<br>6. Do ECD data systems integrate with other sectoral databases (e.g., health, education, child protection)? |
| ECD indicators | Under five | In the past 10 years<br>(01/10/2015 to 30/09/2025) | Academic and grey literature | Regionally, Kenya and county-specific | 7. Which specific ECD indicators are collected in the ECD system (the five areas of NCF – Health, Nutrition, Early Learning, Responsive caregiving, Safety and Security)?<br>8. How frequently is ECD data updated in your system? |
| Challenges specific to ECD data collection and reporting | Under five | In the past 10 years<br>(01/10/2015 to 30/09/2025) | Academic and grey literature | Regionally, Kenya and county-specific | 9. What challenges are being faced in ECD data management?<br>10. What are the primary barriers to data accuracy and reliability? |

**Table 2:** Summary of search strategies per region.

| Region | Search Strategies |
| --- | --- |
| SSA | [("Child" OR "infant" OR "children" OR "ECD") AND ("health*" OR "education" OR "safety" OR "security" OR "protection" OR "stimulation" OR "care*" OR "nutrition" OR "early learning" OR "sanitation" OR "ECD infrastructure" OR "ECD resources" OR "ECD budget allocation" OR "ECD budget expenditure") AND ("data" OR "system" OR "platform" OR "dashboard") AND ("SSA" OR "sub-Saharan Africa") ] |
| Kenya | [("Child" OR "infant" OR "children" OR "ECD") AND ("health*" OR "education" OR "safety" OR "security" OR "protection" OR "stimulation" OR "care*" OR "nutrition" OR "early learning" OR "sanitation" OR "ECD infrastructure" OR "ECD resources" OR "ECD budget allocation" OR "ECD budget expenditure") AND ("data" OR "system" OR "platform") AND ("Kenya")] |
| Nairobi | [("Child" OR "infant" OR "children") AND ("health*" OR "education" OR "safety" OR "security" OR "stimulation" OR "care*" OR "nutrition" OR "early learning" OR "sanitation" OR "ECD infrastructure" OR "ECD resources" OR "ECD budget allocation" OR "ECD budget expenditure") AND ("data" OR "system" OR "platform") AND ("Nairobi")] |
| Isiolo | [("Child" OR "infant" OR "children" OR "ECD") AND ("health*" OR "education" OR "safety" OR "security" OR "protection" OR "stimulation" OR "care*" OR "nutrition" OR "early learning" OR "sanitation" OR "ECD infrastructure" OR "ECD resources" OR "ECD budget allocation" OR "ECD budget expenditure") AND ("data" OR "system" OR "platform") AND ("Isiolo")] |
| Homa Bay | [("Child" OR "infant" OR "children" OR "ECD") AND ("health*" OR "education" OR "safety" OR "security" OR "protection" OR "stimulation" OR "care*" OR "nutrition" OR "early learning" OR "sanitation" OR "ECD infrastructure" OR "ECD resources" OR "ECD budget allocation" OR "ECD budget expenditure") AND ("data" OR "system" OR "platform") AND ("Homa-Bay" OR "Homabay")] |

### Identifying relevant studies

The study team will search for peer-reviewed literature from PubMed, ScienceDirect, Open Grey, African Journals Online (AJOL), Scopus, APA PsycINFO, or PsycNet from October 2015 to September 2025. The grey literature search will utilise online resources, including Google Scholar as a search engine, and national and sub-national websites/reports (such as MoH, social protection, MoE, county governments, NIMEs, NEMIS, and CIMES). The search will include qualitative and quantitative evidence on ECD data systems currently used by various government agencies, as well as the access and challenges encountered while using such systems/platforms.

The literature search strategies for each region are described in Table 2.

**Table 2:** Data extraction form.

|  |
| --- |
| Author |
| Date of publication |
| Title |
| Aims/objectives |
| Settings |
| Population |
| Sampling method |
| Analysis |
| Findings |
| Conclusion |

### Study selection

We will use the following eligibility criteria to select relevant studies to be included in our scoping review:

### Inclusion criteria

All publications from the scientific electronic databases will be eligible for inclusion if they meet the following criteria:

a. Original observational or experimental studies, including quantitative, qualitative, or mixed methods research;
b. Scientific(peer-reviewed) studies or grey literature focusing on children aged 0–5 years;
c. Scientific (peer-reviewed) studies or grey literature covering data collection or collation for any of the components of the nurturing care framework (health, nutrition, opportunities for early learning, responsive caregiving, and safety and security);
d. Published after October 2015 to September 2025
e. Participants from sub-Saharan Africa;
f. Published in English or Kiswahili.

### Exclusion criteria

Studies or grey literature will be excluded based on the following criteria;

a. Systematic/scoping reviews and meta-analyses;
b. Scientific studies or grey literature focusing on children above 5 years only;
c. Published before October 2015;
d. Participants from other regions outside sub-Saharan Africa;
e. Not written in English or Kiswahili.

The eligible studies will first be screened based on the information in their titles and subsequently in the abstracts. Those excluded at this stage will be reported. In the second stage, we will conduct a full-text review to ascertain their eligibility according to the inclusion and exclusion criteria. Finally, each publication will undergo a full-text review to extract all relevant information for the scoping review. The process for study selection is illustrated in Figure 1.

**Figure 1:**
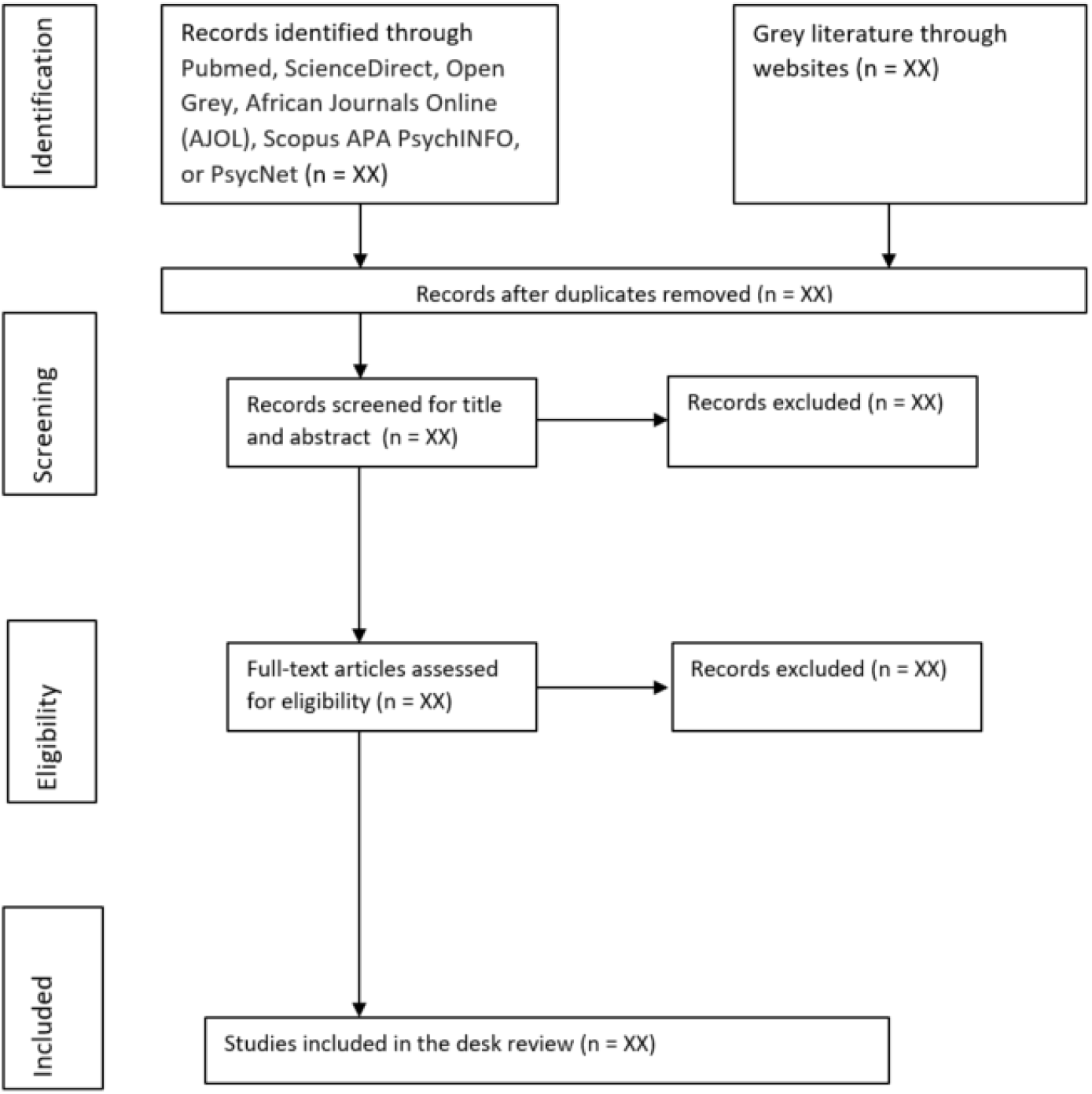
scoping review flowchart.

### Charting the data

We will utilize the data extraction template presented in Table 3 to capture information on each study. The data extraction form will include the authors’ names, publication date, title, study aims/objectives, settings, population, sampling method, analysis, findings, and conclusion. Two reviewers will extract data independently, and discrepancies will be discussed with a third reviewer. Duplicates will be identified across the databases. Studies reported in different sources will be combined during data abstraction.

### Collating, summarizing, and reporting the results

The research team will map evidence and any overall findings associated with the context-specific research questions. Further synthesis will be based on these themes: sources of ECD-related data, types of data systems currently being used, ECD indicators presently being collected, and Challenges specific to ECD data collection and reporting.

### Consultation/stakeholder engagements

The preliminary results of the scoping reviews, including identified potential data visualization systems, will be shared with the three counties’ top leadership for feedback and adjustment where necessary.

## DISCUSSION

This scoping review is expected to play a crucial role in advancing both our immediate research objectives and broader understanding in the field of early childhood development (ECD). First, it will support the next phases of our study by mapping existing ECD data systems in sub-Saharan Africa (SSA), with a particular focus on Kenya. By identifying key stakeholders, indicators, tools, and infrastructure currently in use, the findings will directly inform the creation of an ECD data visualisation platform tailored to the needs of vulnerable communities in Nairobi, Isiolo, and Homa Bay.

Beyond our immediate work, this review will contribute to the wider ECD research and policy landscape by highlighting the fragmentation of ECD information in data systems across SSA. It will identify critical gaps in data collection, management, and utilisation for decision-making throughout SSA. The synthesis of these findings will help pinpoint areas in need of further research, such as the lack of standardised indicators, age-specific data, and real-time monitoring mechanisms. Ultimately, this review will lay the groundwork for evidence-based interventions aimed at improving ECD outcomes in high-risk settings.

### Anticipated Limitations

While this scoping review will provide valuable insights, several limitations must be acknowledged. Firstly, the focus on SSA, especially Kenya, may limit the relevance of findings to other regions with different ECD data systems and challenges. Secondly, the limited number of published studies on ECD data systems in low-resource settings could restrict the review’s depth. Despite these limitations, this review will lay important groundwork for future research and improvements in ECD systems.

## Data Availability

This is a protocol manuscript.

## DISSEMINATION AND ETHICS

### Dissemination

Through our previous and ongoing studies, we have established a strong network with partners in early childhood development and child care across the country. The findings from this review will be used to facilitate a co-creation workshop with stakeholders to design an ECD data visualisation platform. These engagements will involve key stakeholders, including county and sub-county officials, policymakers, local NGOS working in ECD, and child-care centres, who are expected to promote rapid learning, foster buy-in, disseminate study findings, and support the design of the ECD data to enhance the programme’s sustainability.

Furthermore, the staff of the Policy Engagement and Communications Unit at APHRC will facilitate the dissemination of the scoping review report through appropriate media outlets, such as policy briefs, blogs, Twitter, and others. These results will also be shared with the scientific community via publication in peer-reviewed journals and presentations at both local and international conferences.

### Ethics

Ethical approval is not required as this is a review of previously published content. Additionally, this scoping review is part of a study that has received ethical approval from Amref Health Africa’s Ethics and Scientific Review Committee (ESRC), Kenya.

### Authors Contributors

KO led the preparation of the manuscript with inputs from MN. All the authors contributed to the conceptualisation, design and writing of the protocol from which this manuscript was developed. All authors participated in the literature search, selection, and extraction of content from eligible articles, and developed the data extraction tools and methods. All authors (KO, EO, PKW, SO, AA, CW, EM, LO, DM, NL, PO, CWM and MN) participated in the writing, reviewing and editing of the final version of the manuscript.

### Funding

This work is supported by the Conrad Hilton Foundation, Grant Ref Number: 32590

### Patient and public involvement

Patients and/or the public were not involved in the design, conduct, reporting, or dissemination plans of this research.

### Patient consent for publication

Not required.

